# Personalized Prediction of Stress-Induced Blood Pressure Spikes in Real Time from FitBit Data using Artificial Intelligence: A Research Protocol

**DOI:** 10.1101/2023.12.18.23300060

**Authors:** Ali Kargarandehkordi, Peter Washington

## Abstract

**Background:** Referred to as the “silent killer,” elevated blood pressure often goes unnoticed due to the absence of apparent symptoms, resulting in cumulative harm over time. While various health conditions contribute to hypertension, they collectively account for a minority of cases. Chronic stress has been identified as a significant factor in increased blood pressure, and the heterogeneous nature of stress responses makes it challenging to identify specific deleterious behaviors through traditional clinical interviews.

**Objective:** We aim to leverage machine learning algorithms for real-time predictions of stress-induced blood pressure spikes using consumer wearable devices such as FitBit, providing actionable insights to both patients and clinicians to improve diagnostics and enable proactive health monitoring.

**Methods:** The study proposes the development of machine learning algorithms to analyze biosignals obtained from these wearable devices, aiming to make real-time predictions about blood pressure spikes.

**Results:** We have developed the core study application, named CardioMate. CardioMate will be used to remind participants to initiate blood pressure readings using an Omron HeartGuide wearable monitor. The project described is supported as a pilot project from the Robert C. Perry Fund of the Hawai’i Community Foundation. This protocol was approved by the University of Hawai’i Institutional Review Board (IRB) under protocol #2023-00130.

**Conclusions:** Personalized machine learning when applied to biosignals is a promising approach to providing the mobile sensing backend support for real-time digital health interventions for chronic stress and its corresponding symptoms.

## Introduction

### How this Research Benefits the People of Hawai‘i

According to the Department of Health Chronic Disease Prevention & Health Promotion Division, one in every 3 adults in Hawai‘i has been diagnosed with hypertension^1^. Mortality rates associated with heart disease are particularly high for Native Hawaiians and Other Pacific Islanders population, leading to 628 deaths per 100,000 residents as opposed to 154 deaths per 100,000 among Asian and 167 deaths per 100,000 among White residents in Hawai‘i [1].

A recent study conducted by researchers at the John A. Burns School of Medicine found that Native Hawaiians and Other Pacific Islanders under a physician’s care for hypertension experienced an 18.3 point drop in systolic blood pressure on average when participating in a 12-week hula program [2-3]. This study provides strong evidence that stress-reducing interventions can reduce hypertension in Native Hawaiians. We hope to build upon this foundational research by leveraging consumer devices (i.e., a FitBit) to detect moments of high stress and to provide just-in-time interventions which are culturally grounded. The first step of this long-term research plan is to develop the AI which will power the digital intervention, and that first step is the focus of this grant proposal.

### Clinical and Unmet Need

Hypertension is an indirect cause of hundreds of thousands of annual deaths in the United States alone [4]. Known as the “silent killer”[5], elevated blood pressure often remains unnoticed by affected individuals due to lack of perceptible symptoms, resulting in accumulated harm over years. While several causes of hypertension are related to an underlying health condition such as kidney disease, diabetes, sleep apnea, or hormone problems [6], health conditions and medications combined only account for roughly 1 in 20 cases [7]. Chronic stress has been repeatedly documented to increase blood pressure [8-10].

Prior studies have found that elevated blood pressure often arises due to a stressful lifestyle, although the effect of exact stressors varies drastically between individuals. Due to the heterogeneous nature of both the stress and blood pressure response to a multitude of lifestyle decisions, it can be difficult if not impossible to pinpoint the most deleterious behaviors in a personalized manner using the traditional mechanism of clinical interviews. Passive sensing technologies deployed on consumer devices have the potential to disrupt this status quo in a positive manner. By continuously monitoring a patient’s lifestyle in naturalistic settings, digital technologies can provide clinicians and patients alike with actionable insights into their health trends with fine-grained precision.

We are interested in the use of wearable technologies to sense cardiovascular signals, as they are non-invasive and are already widely adopted. We will develop ML algorithms which analyze these biosignals to make real-time predictions about blood pressure spikes. The resulting predictions could be used to alert, in real time, patients about unintentionally adverse behaviors as well clinicians about the frequency of such behaviors. There is a <u>critical opportunity and need</u> to improve diagnostics for repeat health events to enable clinicians to monitor their patients and forecast future issues.

### Innovation

There are countless situations in healthcare and biomedicine where vast amounts of unlabeled data are collected from a single patient [11]. Annotations for the event of interest are frequently sparsely dispersed. The development of predictive supervised machine learning (ML) models is infeasible in such circumstances, as classical approaches cannot handle the complexity of the data and modern deep learning approaches require vast amounts of data [12]. For example, continuous readings from continuously worn glucose monitors can provide enough input data to train a model to make a prediction about patient energy based on glucose, but it is impracticable to require users to log their perceived energy at the same sampling frequency as a wearable device. Similar situations arise from data collected by consumer wearable health devices (e.g., smart watches), smartphones, and other devices which measure biological signals.

To support AI development in these situations where vast longitudinal data are collected with minimal human-provided annotations, we propose the development of ***personalized*** ML models which are trained solely on an individual’s unlabeled data to learn feature representations which are specific to their baseline temporal dynamics. We will train these models with a novel dataset of FitBit biosignals and corresponding blood pressure readings (Figure 1). We are creating a novel method and framework, which has never been explored in healthcare, consisting of pretraining neural networks to learn the temporal dynamics of a patient’s biosignals. This method will enable powerful deep networks to be trained using relatively small datasets which would not be possible without the self-supervised approach proposed here. From a usability standpoint, patients will only be required to provide tens of annotations only tens of times to get a personalized predictive model.

**Figure 1.**
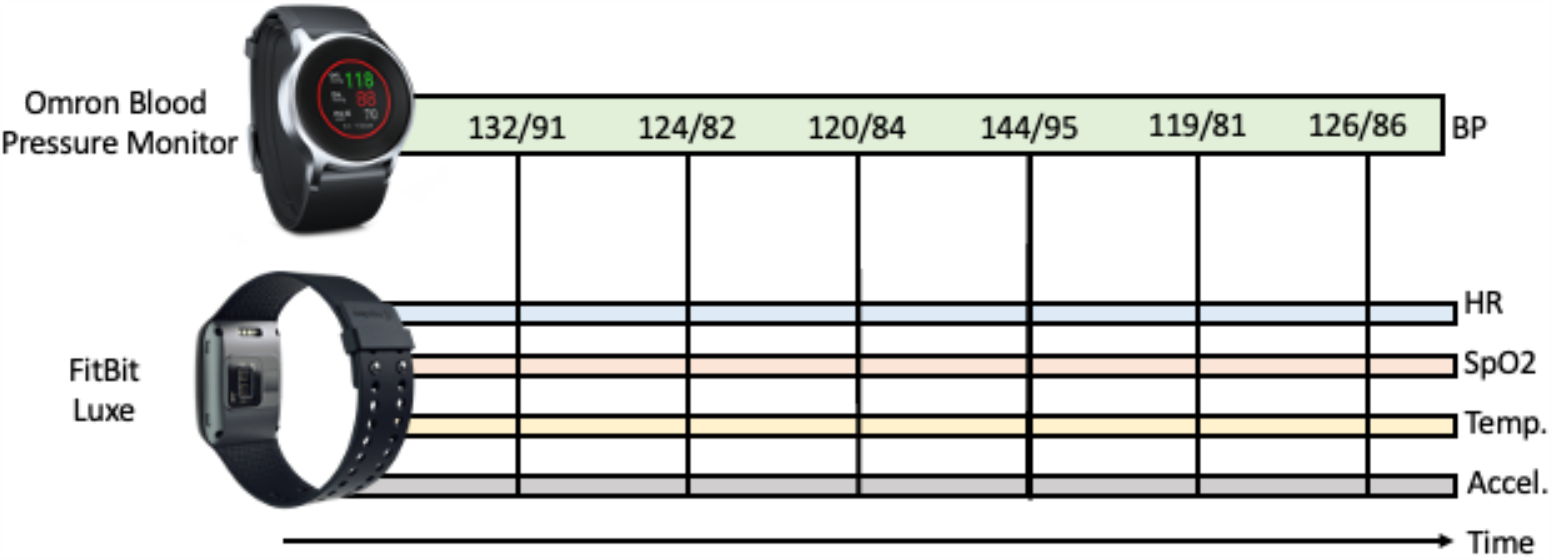
We will collect wearable biosignals from a FitBit and use them to predict blood pressure as measured by an Omron HeartGuide wearable blood pressure monitor. We will use personalized self-supervised learning to enable prediction of blood pressure using minimal samples from the end user.

While we propose to apply this new technological innovation towards the prediction of cardiac signals, multimodal time-series personalization can be applied to a variety of other biology and health problems where (1) multiple signals are emitted, (2) the baseline signal patterns are specific to each individual or organism, and (3) it is infeasible to acquire the vast amounts of labels required to train a supervised deep learning model from scratch. Examples of future applications of the proposed methodology include predictions stemming from Nanopore signal data or multielectrode neuronal recordings. This method has the potential to dramatically advance the field of precision healthcare by enabling reliable ML predictions from massive but mostly unlabeled datasets which are trained in a self-supervised manner on data from a single user.

While this novel methodology could be applied to myriad domains within health and biology, a natural application is the prediction of cardiac events from wearable biosignals data. We will focus on high blood pressure.

### Dissemination Plan

We plan to disseminate our research findings through a combination of (1) research publications in journals, (2) presentations at conferences, (3) as preliminary data for an NIH R01 application, and (4) as the basis of community-based participatory design sessions where we iteratively develop a culturally-informed digital intervention using the AI created in this project. Target journals for submission include Nature Digital Medicine, Science Translational Medicine, IEEE Transactions on Affective Computing, PLoS Digital Medicine, and Cell patterns. Target conferences include the American Medical Informatics Association (AMIA) Annual Symposium, the Pacific Symposium on Biocomputing (PSB), and the Conference for Computer-Human Interaction (CHI). There are several Notices Of Special Interest posted by the NIH which would support a large R01 grant application using the preliminary data from this work.

## Methods

### Specific Aims

We propose the following Specific Aims:

**Aim 1:** Create a novel dataset of wearable sensor data and corresponding blood pressure measurements.

**Aim 2:** Develop a personalized self-supervised pre-training procedure for time series data using both contrastive learning and masked predictions.

**Aim 3:** Develop a novel personalized pre-training procedure which exploits the multimodal nature of wearable time series data.

### Recruitment

We will recruit 40 carefully selected participants with diagnosed hypertension and self-reported stressful lifestyles to each participate in a 4-week remote data collection period. Each participant will wear an Omron HeartGuide blood pressure wearable device and a FitBit Luxe wearable watch during all waking hours for at least 15 hours each day. Apart from wearing the devices and periodically syncing the data to the cloud, participants will be asked to follow their normal routine for the duration of the study.

We will recruit adults ages 30 to 70 in the state of Hawai‘i who have been diagnosed with hypertension and self-identify as living a high-stress lifestyle. Given the diversity of the population of Hawai‘i [13], we aim for the following demographic composition of our participants: ∼23% White, ∼37% Asian, ∼11% Native Hawaiian or Pacific Islander, ∼7% Black or African American, and of ∼22% two or more races. ∼9.5% of the recruited population will have Hispanic or Latino ethnicity.

Dr. Washington has a network of clinical collaborators at the John A. Burns School of Medicine at the University of Hawai‘i at Mānoa who also practice at local medical centers such as Queen’s Medical Center and Kaiser Permanente’s branch in Hawai‘i. I will recruit using the following sources: (1) direct recruitment from the Hawai‘i Pacific Health Center, which my collaborators at the University of Hawai‘i Department of Psychiatry are affiliated with and where they practice clinically, (2) via flyers and emails at the clinics which the University of Hawai‘i Department of Psychiatry regularly provides inpatient and outpatient psychiatric services and consultation at: including The Queen’s Medical Center, Kapi‘olani Medical Center for Women and Children, and Hawai’i State Hospital Community mental health centers on Hawai’i Island, Moloka‘i, Maui, Kaua‘i, and Lāna‘i, (3) advertisements posted on the University of Hawai‘i campus and in public settings in Honolulu, and (4) targeted advertisements posted to social media websites. I will work with Dr. Guerrero, the chair of the Department of Psychiatry at the University of Hawai‘i, to ensure recruitment strategies and advertisement of the research program translates across cultures and to ensure effective recruitment as well as diverse and representative data.

We will exclude participants younger than 30 years and older than 70 years of age. We will require all potential participants to remotely complete the Perceived Stress Scale (PSS), a 10-item scale which is the most widely used psychological instrument for measuring the perception of stress [14]. We will exclude participants whose PSS score does not exceed one standard deviation above the mean for at least one of their demographic brackets (age, gender, or race) as reported by Cohen et al [14].

We will also ask participants to self-report their blood pressure. We will remotely ask participants whether they are currently taking any blood pressure medication, and we will exclude all such participants. We will also exclude participants who do not own a smartphone with continuous network connectivity. During the in-person study intake, we will measure blood pressure of potential study participants three times. We will exclude participants whose blood pressure does not exceed 130/80 mmHg for at least one of the measurements, as 130/80 mmHg is the minimum cutoff for Stage 1 hypertension.

### Data Collection and Storage

We will leverage the existing application programming interface (API) provided by both Omron and FitBit to record the user’s wearable sensor readings and upload the data to the cloud. Omron’s Healthcare API offers access to timestamped blood pressure readings as well as activity and sleep approximations. The FitBit API provides access to sensor readings of heart rate (HR), gyroscope, accelerometer, breathing rate, blood oxygen levels (SpO2), and skin temperature sensor readings.

The data will be managed on each participant’s smartphone devices through an application, implemented for both iOS and Android, that we will develop. The study team will install the application on the user’s smartphone and configure the Omron and FitBit device during study onboarding. The smartphone application will periodically trigger a notification reminding the participant to (1) measure their blood pressure with the Omron wearable, (2) sync the Omron and FitBit data to the application, and (3) connect to a network while the study app is open to allow the data to be uploaded to a centralized server.

We will store the curated data from each participant on a centralized server hosted on Amazon Web Services (AWS). Because FitBit is owned by Google, participants’ FitBit data will be uploaded directly to Google’s cloud servers, which utilizes the same level of security as other Google products such as Gmail. Access to each participant’s FitBit data on Google’s cloud servers is implemented through OAuth, which provides clients with a secure delegated access to server resources on behalf of a resource owner (i.e., the participants of this study). This mechanism is used by companies such as Amazon, Google, Facebook, Microsoft, and Twitter to permit the users to share information about their accounts with third-party applications or websites. In this case, the “third party” is the study team. The FitBit data and blood pressure readings will be preprocessed on an Elastic Cloud Compute (EC2) instance on AWS, which is HIPAA-compliant. The EC2 instance will store the data onto respective database tables using DynamoDB. Each table will have columns for the child ID and the timestamp. We will encrypt all server-side data and require secret access keys for data access. DynamoDB tables are automatically encrypted on the server side. To add an additional layer of security, we will implement client-side encryption on the mobile application, ensuring encrypted data transmission over an HTTPS connection to move blood pressure data between the devices and AWS. Data access will require a secret access key provided by the AWS administrators to any data analysis team. The data will not be accessible without this key. For further security, we will anonymize all user data on the server side by removing all PHI from the DynamoDB tables.

We intend to release the curated data (Figure 2) as a publicly available dataset for use in the evaluation of multimodal time series ML models. Such datasets exist for activity and emotion recognition from wearable data, but prediction of blood pressure from these measurements will be a challenging task that other researchers can attempt with the release of our dataset. This will be the first publicly available dataset which includes at-home blood pressure measurements alongside wearable sensors like HR, SpO2, and accelerometer readings. This fully anonymized dataset will only be released to researchers who sign a Data Use Agreement which will be approved by the University of Hawai‘i Data Governance Office.

**Figure 2.**
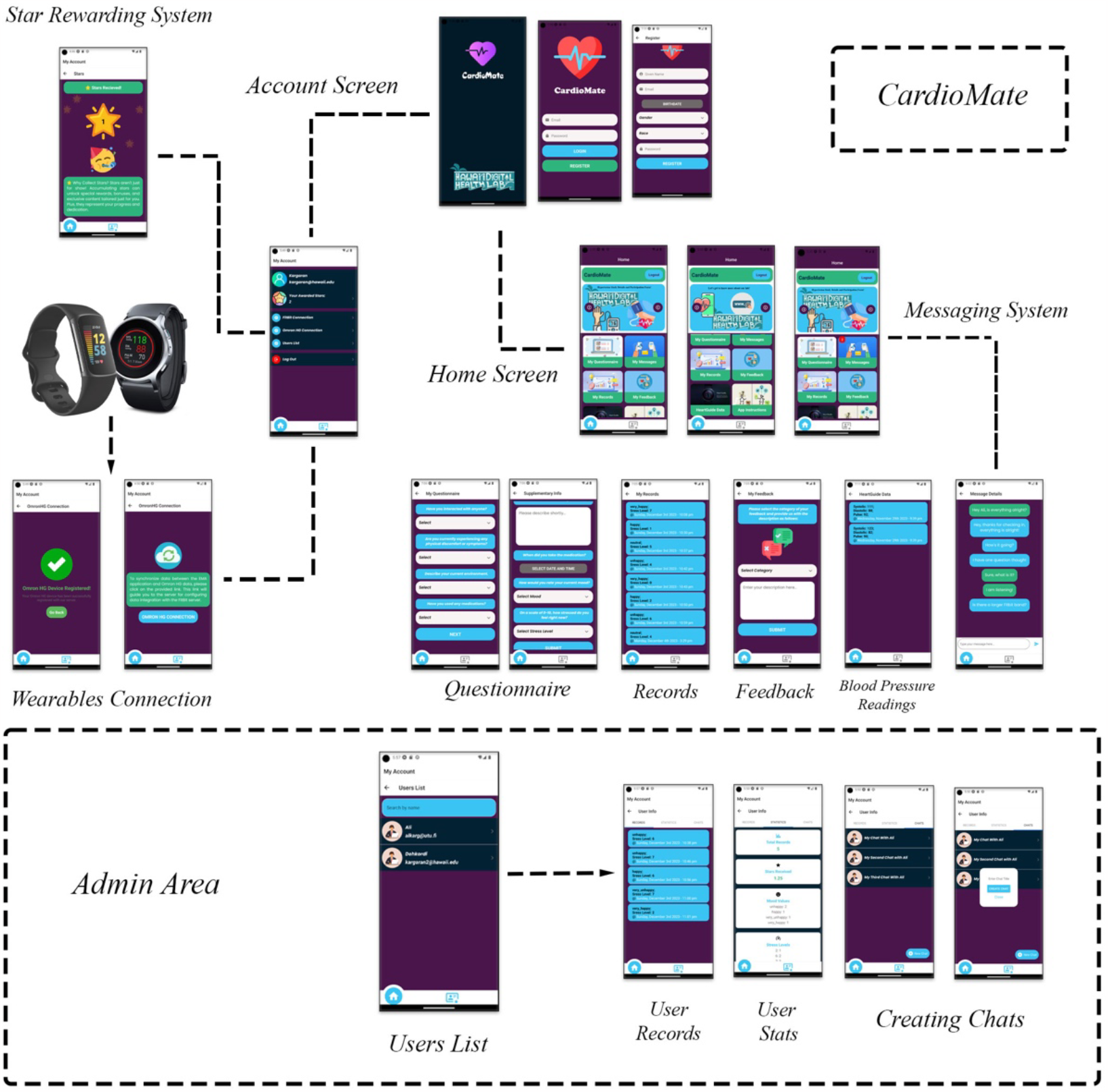
Workflow of the CardioMate app. The application comprises two primary screens: Account and Home. The Account screen features user details, a star reward system for active participation in the study, and options to link two wearable devices (Fitbit and Omron Heartguide) for data synchronization with our secure, encrypted database. The Home screen is divided into six sections, including Questionnaire, Messages, Feedback, Records, Blood Pressure Readings, and App Instructions. Additionally, the CardioMate app includes an administrative area for study managers to view participant statistics and initiate personalized chats, complete with alarm and notification functions.

### Feasibility

The most difficult aspect of this Aim will be maintaining participant engagement throughout the 4-week study period. The graduate research assistant funded by this project will dedicate some time each day towards running the study and interfacing with participants. We expect participants to open the smartphone application to sync and upload their data on a daily basis, which is a 1-minute time commitment per day.

While we expect no trouble recruiting 40 subjects for participant, we expect some participants to drop off during the study. Since we will have enough devices for 5 concurrent subjects, it will take 8 months to collect all data if no participants drop off. Our study timeline allocates 6 additional months of make-up time to collect data from new participants, accounting for >50% drop-off rate. Given the remote nature of the data collection procedures, we expect some participants to drop off from the study prematurely or to not comply with study processes. We will therefore remotely monitor the upload progress and send an automated text and email notification to the participant if the data were not uploaded in a timely manner. If 3 consecutive days of participant non-compliance are detected, we will contact the participant for device return.

### AI Model Training

SSL is usually used to pre-train an entire dataset with no explicit labeling by humans to guide the supervision task. We propose to redesign the SSL paradigm towards the task of model personalization. By pre-training a model only on the vast amounts of data curated from a single individual, the weights of the neural network will learn to make predictions using the inherent structure of each participant’s biosignals. This is essential because baseline HR, SpO2, skin temperature, and movement patterns, regardless of stress, will vary drastically across individuals, limiting the performance of general-purpose ML models.

To train ML models which predict blood pressure based on a user’s wearable biometrics, we will develop and evaluate a series of both long short-term memory (LSTM) and Transformer neural networks. The inputs to the models will consist of a separate 1D convolutional backbone for each biometric modality. The convolutional features will be fused upstream into a shared joint dense representation space and finally a dense prediction layer with linear activation for regression prediction. We will implement all models using Tensorflow [15].

We will perform a series of self-supervised pretraining tasks to allow the networks to learn the baseline temporal dynamics of each individual’s biosignals. As one pretraining task, we will develop contrastive learning methods to automatically learn embeddings which encode the structure of the signal. For each wearable sensor modality, we will run a sliding window to isolate short time segments. We will apply signal-based data augmentation techniques to derive a new signal. We will perform contrastive learning to learn neural network embeddings which maximize the similarity between each original segment and its modified version while minimizing the similarity across segments (Figure 3).

We will develop a modified version of the SimCLR algorithm which will be tuned for the task of personalization to a user’s wearable signal readings. It is often the case that biosignals look highly similar, either due to temporal locality or by relative homogeneity of the individual’s activity. To account for this possibility of recurring signal patterns, we will weight the attract and repel strength of SimCLR based on the temporal distance between two segments of a particular signal. We will run grid search to tune this repel strength.

The data augmentation techniques that we apply to the signals will be domain-specific, keeping in mind the inherent nature of each sensor. For example, for accelerometer data, rotations simulate different sensor placements and cropping is used to diminish the dependency of event locations [16]. Across several modalities, sensor noise can be simulated through scaling, magnitude-warping, and jittering [16]. We will be careful to not apply augmentation strategies which might change the meaning of the underlying signal.

As another pre-training task, we will perform generative pre-training by masking the input signal and predicting the missing portion of the signal using a deep autoencoder architecture (Figure 4). Pretraining in this manner will teach the model to understand the dynamics of each time series signal independent of blood pressure or any other labels.

We will train the model on the first 60% of data (by time), tune hyperparameters on the next 20% of data, and calculate the mean absolute error (MAE) and mean squared error (MSE) on the final 20%. This evaluation pattern mimics real-world use, where a model will be calibrated by a user prior to real-world deployment. It is important to emphasize that we will train and test a separate personalized ML model for each individual.

We will evaluate the models by comparing the performance with respect to the number of labeled examples used for supervised fine-tuning. A plot of this comparison will elucidate the number of blood pressure measurements required for model calibration to a single individual. We will plot the MSE at 10, 20, 30, 40, 50, 75, 100, 125, and 150 blood pressure annotations, as these are feasible amounts of labels that might be provided by a user in real-world use. To ensure a robust evaluation, we will bootstrap at least 20 random samples of blood pressure annotation subsets for each point on the x-axis and will report the mean and 90% confidence interval. Just as in the plain supervised learning condition, we will create a separate plot for each study participant, as the ML portion of this proposal is testing the personalization of ML models rather than a general-purpose one-size-fits-all ML model which is more typical in ML evaluations.

We will perform a similar style of analysis for other clinical outcomes using publicly available datasets such as the Wearable Stress and Affect Detection (WESAD) [17] dataset, a multimodal sensor dataset for stress detection of nurses in a hospital [18], and K-EmoCon, a multimodal sensor dataset for continuous emotion recognition in naturalistic conversations [19]. Each of these datasets, as well as several other publicly available datasets, contains several hours of multimodal biosignal data that overlap with the signals that we propose to collect, such as skin temperature, accelerometer streams, and heart rate. These datasets also include timestamped annotations of endpoints that are likely to be correlated with blood pressure, including self-perceived stress.

In prior work by other researchers, SSL pre-training approaches have repeatedly demonstrated improved performance over pure supervised learning in a variety of contexts [20-23]. Our preliminary data (see Results section below) support that self-supervised pre-training on data solely from each individual results in improved models over pure supervised learning. While unlikely given our preliminary data and prior SSL publications, it is possible that minimal performance gains will be observed when applying the SSL strategies in a personalized manner. In such cases, the negative result would be a noteworthy finding due to prior successes of SSL.

## Results

We have developed a smartphone application, CardioMate, that will prompt participants to measure their BP and log their stress (Figure 2). The application comprises two primary screens: Account and Home. The Account screen features user details, a star reward system for active participation in the study, and options to link two wearable devices (Fitbit and Omron Heartguide) for data synchronization with our secure, encrypted database. The Home screen is divided into six sections, including Questionnaire, Messages, Feedback, Records, Blood Pressure Readings, and App Instructions. Additionally, the CardioMate app includes an administrative area for study managers to view participant statistics and initiate personalized chats, complete with alarm and notification functions.

Our initial sets of published experiments have demonstrated promise for personalized SSL of stress but with some caveats. Our experiments on the WESAD dataset demonstrated that deep learning model performance improves drastically when using self-supervised personalization when compared to personalization without SSL when there are a small number of labeled data points for supervision [24]. This effect diminishes with increasing amounts of labeled data [25-26], aligning with prior work that demonstrates that SSL is only beneficial under low-label settings. We have also tried these methods on a particularly challenging dataset: a multimodal sensor dataset for stress detection of nurses in a hospital [18]. This dataset consists of wearable biosignals measured from nurses who wore Empatica E4 wristbands while conducting their normal shifts. This dataset is difficult because (1) the data were collected in the wild rather than in controlled lab settings and (2) individual nurses were not consistent about their labeling practices, leading to sparse, irregular, noisy, and otherwise messy labels. Consequently, we found that the difference in AUC-ROC scores for self-supervised models was only about 2.5% higher on average compared against an equivalent baseline model [27], and this increase is within the margin of error due to the limited sample size. This lack of improvement in noisy annotation settings highlights the need for HCI innovations for improving data labeling quality for personalized AI within naturalistic settings.

We have also observed improved performance when personalizing affect-related prediction tasks without personalization both using classical ML [28] and deep learning [29] as well as when only applying SSL without personalization [30]. When disentangling and comparing the effects of SSL and personalization separately, we find that SSL yields more benefit than individualization on non-affective medical data with large time intervals between data points, suggesting that the sampling frequency and other data considerations must be considered [31]. Collectively, these preliminary results demonstrate promise for the core ML approach that we propose.

## Data Availability

All data produced in the present study are available upon reasonable request to the authors

## Acknowledgements

This project received a grant (#MedRes_2023_00002689) from the Robert C. Perry Fund of the Hawai’i Community Foundation.

